# Use of a Single Case-Finding Questionnaire to Simultaneously Target Multiple Related Diseases Allows Enhanced Disease Detection

**DOI:** 10.1101/2023.12.08.23299740

**Authors:** Shawn D. Aaron, Chau Huynh, George Alex Whitmore

## Abstract

**Objective:** To develop a research methodology to apply a single case-finding tool to multiple related diseases and to evaluate the ability of a single tool to detect two or more related chronic diseases.

**Methods:** Adults in the community with no prior history of physician-diagnosed lung disease who self-reported respiratory symptoms were contacted via random-digit dialing. Multiple risk scores, one for asthma and one for COPD, were developed using data from a single case-finding questionnaire administered to the study population. Each score was statistically optimized for targeted detection of cases having one disease in the class. External validation of tandem risk scores was prospectively conducted in an independent sample and predictive performance re-evaluated.

**Results:** Sensitivity for detection of asthma improved from 87% using single risk scores to 96% using tandem risk scores, and sensitivity for detection of COPD similarly improved from 87% to 99%. In the independent validation cohort, case-finding sensitivities increased from 64% and 59% using single risk scores to 95% and 96% using tandem risk scores for asthma and for COPD, respectively.

**Conclusions:** Use of a single questionnaire which incorporates risk scores for multiple diseases considered in tandem, rather than individually, enhances the yield of cases detected when compared with one-at-a-time application of risk scores for case discovery. Benefits include greater efficiency in case-finding and improved sensitivities for detection of each disease.

**What is New?:**

- We describe case finding in a population of undiagnosed, symptomatic subjects who have one disease from a class of related chronic diseases.
- Multiple risk scores are developed using data from a single case-finding instrument administered to a representative sample from the study population. Each score is statistically optimized for targeted detection of cases having one disease in the class.
- Use of multiple risk scores, when considered in tandem, enhances the yield of cases detected when compared with one-at-a-time application of risk scores for case discovery.

## Introduction

Some chronic diseases or syndromes may have broad similarities or may overlap in their presentations. These related diseases may present with similar symptoms, have common negative health outcomes, or share similar pathophysiologic mechanisms. Examples of related diseases include asthma and chronic obstructive pulmonary disease (COPD) which present with similar symptoms (dyspnea, cough, wheeze and/or chest tightness) and which share expiratory airflow obstruction as a common physiologic impairment. Other examples of related diseases which share similar symptoms include ulcerative colitis and Crohn’s disease, or cardiomyopathy and valvular heart disease.

Since these chronic diseases are relatively prevalent in populations, it might be of interest for clinical researchers to conduct case finding studies to detect subjects in the community who are suffering symptoms of these individual chronic diseases but who remain undiagnosed [1]. In the case of related diseases, it is often more cost-effective to detect subjects (discover cases) having one of multiple related diseases based on a single case-finding questionnaire or instrument. In deriving and testing the instrument, subjects would complete the questionnaire and then subsequently undergo gold standard diagnostic assessments for disease. Using this approach, it is possible to develop and calculate a different risk score formula for each individual disease from a common survey instrument.

Our objective was to develop a research methodology to apply a single case-finding tool to multiple related diseases and to evaluate the ability of a single tool to detect two or more related chronic diseases. We demonstrate that a benefit of this concurrent detection strategy is higher predictive performance from considering risk scores jointly (i.e., in tandem). The false negative rates across all diseases can be lower in aggregate if the diseases are considered in tandem rather than if each is considered separately, and thus the total social and economic costs of prediction errors (missed cases) can be lowered significantly.

## Methods

Our method considers a case-finding program with the following design elements:

1. <u>Identification of a study population of eligible subjects</u>. The study population consists of at-risk individuals who may have one disease from a class of multiple distinct but related diseases. The study population is at risk of disease by virtue of symptoms or exposure to disease-causing elements (ex. smoking in the case of COPD).
2. <u>Administration of multiple health status and demographic questionnaires to at-risk subjects sampled from the study population</u>. This sample serves as a *derivation sample* for developing the single case-finding questionnaire and individual disease risk score formulas.
3. <u>Determination of a diagnosis</u>. Upon completing step 2, subjects undergo definitive diagnostic evaluation to determine which disease in the class they may have, if any.
4. <u>Development of a single case-finding questionnaire and multiple risk score formulas</u>. With the data record and clinical diagnosis in hand for each subject, multinomial logistic regression or a similar statistical technique is used to develop risk score formulas that relate each subject’s diagnosis to all relevant descriptive data and question responses from the subject’s record. A single common case-finding questionnaire is developed from these data. In the situation of two related diseases, the statistical analysis produces two risk scores, based on the subjects’ responses to the case-finding questionnaire, that are each optimized for one of the diseases.
5. <u>Setting case discovery thresholds</u>. The investigative team decides on a threshold or cut point for each risk score which, if exceeded, identifies the subject as a potential case. Letting t1 and t2 denote the respective thresholds for risk scores R1 and R2, the decision rule for identifying a potential case is finding R1 ≥ t1 and/or R2 ≥ t2. The thresholds are chosen so they strike the best balance for risks of false positive and false negative outcomes.
6. <u>Validation of risk scoring formulas and thresholds</u>. The proposed method for case finding must demonstrate that it is effective when using an independent *validation sample*. Once validated, the method may be implemented in clinical case-finding projects.

### Case Illustration-The UCAP Study: Finding cases of asthma and COPD

We demonstrate the implementation of our enhanced case-finding methodology by reporting results for a prospective study called The Undiagnosed COPD and Asthma Population (UCAP) study, which is aimed at detecting obstructive lung disease cases (either asthma or COPD) among symptomatic undiagnosed adults in the general Canadian population [3, 4]. Eligible subjects with respiratory symptoms who had no history of diagnosed respiratory disease were invited to local study sites for pre-and post-bronchodilator spirometry to confirm, or rule out, asthma or COPD based on their spirometry. The study was recruited participants from June 19, 2017 until March 10, 2020. All participating individuals signed informed written consent. The study received ethical approval from the Ottawa Health Science Network Research Ethics Board (protocol 2017012-01H). The data set contains 1580 subjects, of whom 135 were ultimately diagnosed with asthma and 190 diagnosed with COPD.

Study participants completed a set of five health-status and symptom questionnaires used in respiratory research as well as personal data questionnaires covering demographic and clinical topics. Multinomial logistic functions were derived by regressing subjects’ disease outcomes on subjects’ responses to questions from the survey instruments [5]. Stepwise variable selection was employed to choose a small set of variables for the final case-finding questionnaire and risk score formulas [4]. A single thirteen-item case-finding questionnaire (named the UCAP Questionnaire or UCAP-Q) was developed, and two risk score formulas were derived; one risk score was optimized for detecting asthma and the other for COPD [4]. The risk scores used in this demonstration are the predicted probabilities (in %) of asthma (R1) and COPD (R2) calculated from the fitted multinomial logistic model. A public-use UCAP Questionnaire online calculator for these risk scores can be found at https://omc.ohri.ca/UCAPquestionnaire/.

Threshold risk probabilities for either disease (t1 and t2) were chosen based on the relatively large social costs of missing true cases of either asthma or COPD. Thus, for the purposes of this example our goal was to maximize sensitivity (ability of the questionnaire to detect new cases of disease) at the expense of specificity. In this report, we present illustrative results for thresholds fixed at t1=5% probability for asthma and t2=7% probability for COPD. We choose different thresholds for the two diseases to demonstrate the generality of the method.

#### Classification table for the derivation sample

The classification table for case-finding results based on a single risk score for a single target disease is a conventional 2 × 2 table with two predicted outcomes (positive prediction, negative prediction) and two true disease states (no-disease, disease) [2]. With multiple diseases, the classification table expands in size accordingly. For two diseases, the classification table takes the general form shown in **Table 1**, which illustrates asthma and COPD case finding results in 1580 subjects from the derivation sample of the UCAP study. Rows 1 to 3 of the table show the different risk-score patterns which lead to predicting that a subject has one of the two diseases (a positive prediction). Row 4 gives the numbers of subjects for whom risk scores R1 and R2 lie below their respective thresholds t1 and t2, thereby leading to a prediction of no disease (a negative prediction). Row 5 shows the total number of subjects in each true disease state.

**Table 1:** Case-finding classification table for tandem risk analysis of two related obstructive lung diseases for the risk-score derivation sample.

|  |  | True Disease State |  |  |  |
| --- | --- | --- | --- | --- | --- |
| Column | 1 | 2 | 3 | 4 | 5 |
| Row | Disease Prediction | No Obstructive Lung Disease | Asthma | COPD | Total |
| 1 | Predict Asthma<br>$R1 \geq t1, R2 < t2$ | 574 | 76 | 22 | 672 |
| 2 | Predict COPD<br>$R1 < t1, R2 \geq t2$ | 191 | 13 | 95 | 299 |
| 3 | Predict both Asthma and COPD<br>$R1 \geq t1, R2 \geq t2$ | 330 | 41 | 71 | 442 |
| 4 | Predict No Disease<br>$R1 < t1, R2 < t2$ | 160 | 5 | 2 | 167 |
| 5 | Total | 1255 | 135 | 190 | N=1580 |
| 6 | Sensitivity<br>(Single Risk Scores) | - | 87% | 87% |  |
| 7 | Sensitivity<br>(Tandem Risk Scores) | - | 96% | 99% |  |
| 8 | Specificity | 13% | - | - | - |

Table 1 illustrates the cases that are discovered by a traditional case finding strategy targeting a single disease. For asthma, the cells in column 3 with counts of 76 and 41 subjects are 117 asthma cases that would be discovered using risk score R1 because R1 ≥ t1 for each of these subject groups. Similarly, if the case-finding investigation were targeting only COPD using the single risk score R2, the cells in column 4 with counts of 95 and 71 subjects are 166 COPD cases that would be discovered because R2 ≥ t2 for each of these subject groups.

Next, the table also shows the case-finding gains offered by targeting both diseases in tandem. These gains are found in the two yellow-shaded cells in the table having counts of 13 and 22. These two cells have the distinguishing characteristic that they count subjects whose risk score for only one disease exceeds its threshold, but the subject is discovered to have the other disease. For example, the cell with 22 subjects is counting subjects who were predicted by the UCAP questionnaire to have asthma but in truth have COPD. Similarly, the cell with 13 subjects is counting subjects who were predicted by the questionnaire to have COPD, but in truth have asthma. We refer to these subjects as *cross-over* cases. These cross-over subjects would not be detected if the case-finding investigation is limited to detection of only one of the two diseases using one risk score. Hence, our method of case finding for multiple related diseases increased detected cases by 12%, lifting detection totals by 35 cases, from 283 cases to 318.

The enhancement of case finding for the derivation sample is also demonstrated by comparing the sensitivity values achieved with the tandem and conventional case detection methods. Calculations from Table 1 (rows 6 and 7) show that sensitivity for detection of asthma is improved from 87% using single risk scores to 96% using tandem risk scores, and that sensitivity for detection of COPD is improved from 87% using single risk scores to 99% using tandem risk scores. Row 8 in the table reports the single specificity value of 13% for the tandem case detection method.

#### Classification table for the validation sample

Table 2 presents the cross-classification table for an independent, prospective validation sample of 471 subjects. The cross-over case numbers are again reported in the yellow-shaded cells. In this validation sample, a total of 56 cases would have been discovered using single risk scores, as can be seen by adding the unshaded cells for each disease (27 asthma cases and 29 COPD cases in total). The cross-over cases are found to total 31, consisting of 13 asthma cases and 18 COPD cases. The cross-over cases have therefore increased the total number of cases discovered in the validation sample from 56 to 87. This increase amounts to a 55% improvement in case finding. Consequently, case-finding sensitivities increase from 64% using single risk scores to 95% using tandem risk scores for asthma, and from 59% to 96% for COPD.

**Table 2:**
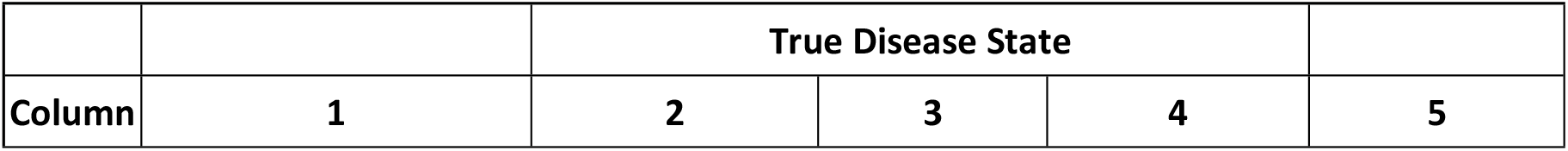
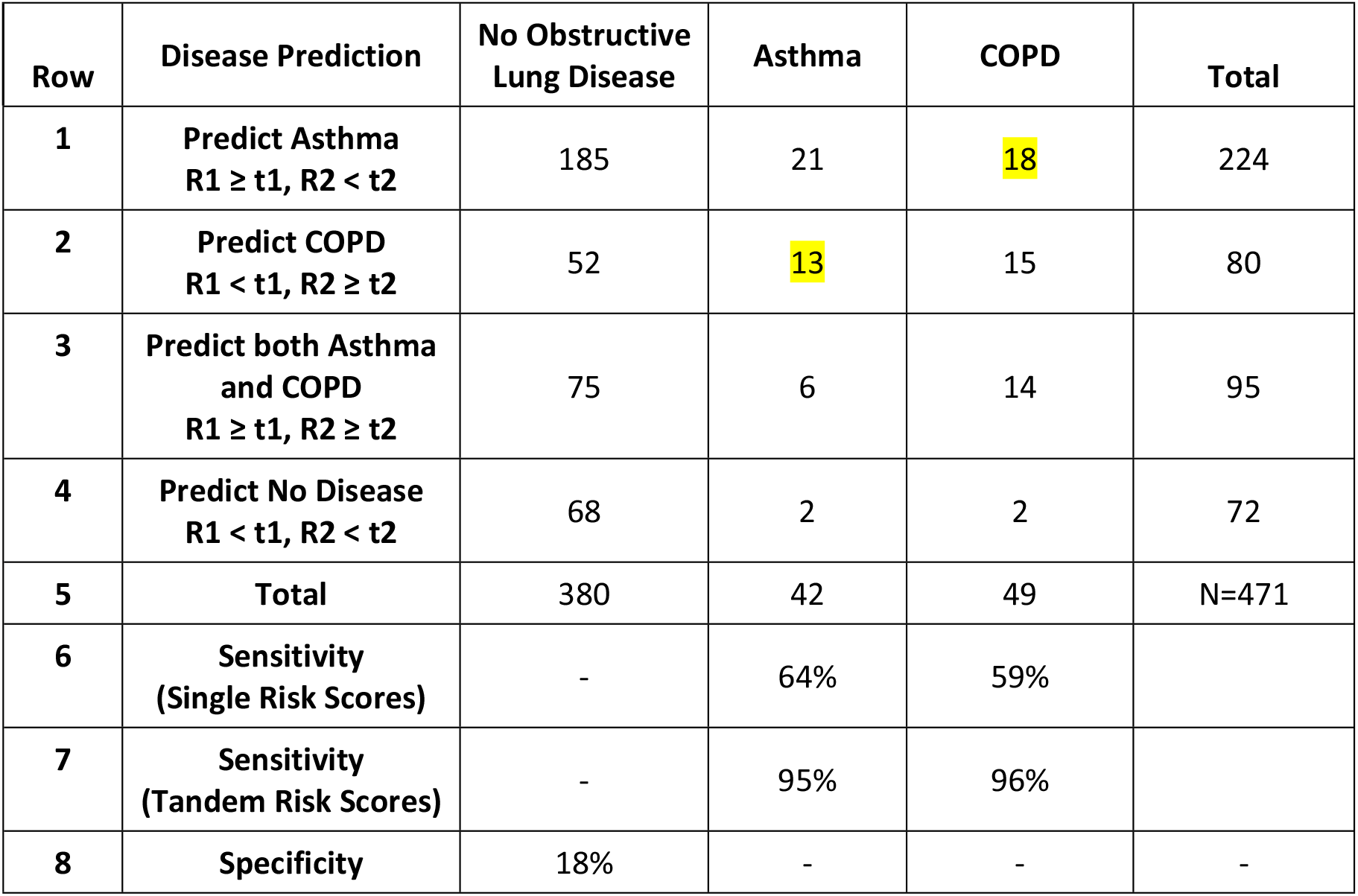
Case-finding classification table for tandem risk analysis of two related obstructive lung diseases for the risk-score validation sample.

## Discussion

Our case illustration has involved creating two risk scores for two related diseases from a single case-finding instrument. An extension to more than two related diseases does not require new concepts, principles or methods. Having more diseases simply increases the size of the classification table. An example of a three-disease setting, where the ‘diseases’ are three severity stages for cancer (mild, moderate and severe) is described by Berger et al [6].

The benefit of concurrent case detection for related diseases can be substantial, both in terms of case discovery and clinical understanding. The cross-over subjects represent a pure gain in case detection at little extra cost. The costs only involve the calculation of a second risk score (using available data from the case-finding questionnaire) and an extended clinical evaluation for the presence of the second disease. The cross-over subjects also offer potentially valuable lessons for disease diagnosis as these subjects have clinical presentations (as determined from their responses on the case-finding questionnaire) that appear inconsistent with their actual disease. Our results for the case illustration show that cross-over cases have the clinical presentation of the ‘other’ disease. Asthma cross-over cases have the clinical appearance of COPD and vice versa. In this respect, these cross-over cases are significant if they represent overlooked or misdiagnosed cases. Statistical methods can be used to probe the apparent dissonance in these cross-over cases.

Our construction of tandem risk scores depends on the predictive information provided by a single case-finding instrument. Understanding the phenomenon of cross over and clinical overlap in disease presentations is critical for selecting and/or creating good questions for the case finding instrument. As always, pilot questionnaire testing, and other good survey practices, are important.

## Conclusion

When a single case-finding questionnaire is used in case finding for a class of related diseases, it is reasonable to develop a separate risk scoring formula for each disease in the class. Our case illustration shows that considering the risk scores in tandem rather than separately enhances the yield of detected cases. We show, not surprisingly, that the extra yield comes from cases that tend to be similar to other diseases in the class and, hence, might be missed in traditional single-disease case-finding studies. Tandem risk analysis shows gains in sensitivity values for our case illustration, which may also be the experience for case finding with other classes of related chronic diseases. These gains can be large in terms of numbers and types of cases found, and generally would be achieved with little extra operational cost.

## Data Availability

All relevant data are within the manuscript and its Supporting Information files.

## Notes

### Competing Interest Statement

The authors have declared no competing interest.

### Clinical Trial

NA

### Funding Statement

Yes

### Author Declarations

The study obtained approval from the Ottawa Health Science Network Research Ethics Board and Clinical Trials Ontario (CTO). All participants provided informed, written consent. CTO Project ID: 1357 SHORT TITLE: UNDIAGNOSED COPD AND ASTHMA POPULATION STUDY (UCAP) Short Study Title: UCAP {UNDIAGNOSED COPD AND ASTHMA POPULATION STUDY} Sponsor Study ID (if applicable): REB of Record: OHSN-REB (General Campus Panel)

